# Evidence of compensatory neural hyperactivity in a subgroup of chemotherapy-treated breast cancer survivors and its association with brain aging

**DOI:** 10.1101/2024.04.22.24306190

**Authors:** Michele M. Mulholland, Alexa Stuifbergen, Alexa De La Torre Schutz, Oscar Y. Franco Rocha, Douglas W. Blayney, Shelli R. Kesler

## Abstract

Chemotherapy-related cognitive impairment (CRCI) remains poorly understood in terms of the mechanisms of cognitive decline. Neural hyperactivity has been reported on average in cancer survivors, but it is unclear which patients demonstrate this neurophenotype, limiting precision medicine in this population. We evaluated a retrospective sample of 80 breast cancer survivors and 80 non-cancer controls, age 35-73, for which we had previously identified and validated three data-driven, biological subgroups (biotypes) of CRCI. We measured neural activity using the z-normalized percent amplitude of fluctuation from resting state functional magnetic resonance imaging (MRI). We tested established, quantitative criteria to determine if hyperactivity can accurately be considered compensatory. We also calculated brain age gap by applying a previously validated algorithm to anatomic MRI. We found that neural activity differed across the three CRCI biotypes and controls (F = 13.5, p < 0.001), with Biotype 2 demonstrating significant hyperactivity compared to the other groups (p < 0.004, corrected), primarily in prefrontal regions. Alternatively, Biotypes 1 and 3 demonstrated significant hypoactivity (p < 0.02, corrected). Hyperactivity in Biotype 2 met several of the criteria to be considered compensatory. However, we also found a positive relationship between neural activity and brain age gap in these patients (r = 0.45, p = 0.042). Our results indicated that neural hyperactivity is specific to a subgroup of breast cancer survivors and, while it seems to support preserved cognitive function, it could also increase the risk of accelerated brain aging. These findings could inform future neuromodulatory interventions with respect to the risks and benefits of up or downregulation of neural activity.

## Introduction

Chemotherapy-related cognitive impairment (CRCI) is experienced by many patients during and after cancer treatment. Despite affecting up to 85% of cancer survivors (1), CRCI is still poorly understood. Clinical and preclinical research from our group and others suggests that breast cancer chemotherapy upregulates neural activity (2–7). Although hypoactivity compared to non-cancer controls has also been observed (8, 9), hyperactivity is more common, especially longitudinally, and is correlated with subjective cognitive function (6, 7, 10, 11). Hyperactivity is not limited to breast cancer. For example, Liu et al. found that colorectal cancer patients treated with chemotherapy had greater activation in several brain regions compared to healthy controls. However, it is unknown which patients show neural hyperactivity as most observations have been made by comparing mean activity between patients and controls. It is possible that a specific subgroup of patients demonstrate hyperactivity, contributing to the heterogeneity in findings across imaging studies.

To identify CRCI subgroups, we pioneered the application of biotyping to this population (12–14). Specifically, we developed an AI-based algorithm for determining data-driven, latent patterns of brain abnormality (biotypes) in breast cancer survivors. We then examined cognitive phenotypes associated with each biotype (14). As we previously described, Biotype 1 demonstrated impaired cognitive function, Biotype 2 had relatively preserved cognitive function, and Biotype 3 showed moderately impaired cognitive function. Impairment was defined as differing significantly from non-cancer controls, although biotypes also differed significantly from each other. We then cross-validated our biotype algorithm in an independent sample and showed that biotypes had unique demographic, clinical, psychological, and genetic characteristics. In contrast, traditional, symptom-based definitions of cognitive impairment showed no significant differences in these characteristics (12, 14). In the present study, we hypothesized that Biotype 2 would uniquely demonstrate neural hyperactivity given their relatively preserved cognitive function.

The basis for this hypothesis stems from research suggesting that neural hyperactivity can be compensatory, or a reorganization of brain function to counteract decline (15, 16). In CRCI studies, hyperactivity is often interpreted as compensatory without any evidence to support this claim (5, 17–20). Cabeza and Dennis (21) proposed four criteria that researchers could use to determine if brain activity can be attributed to compensation (Figure 1). The first two criteria describe “attempted compensation”, indicating that hyperactivity has an inverted U-shaped relationship with brain decline, task demands, and age. Criterion A indicates that there is an initial increase in brain activity related to brain decline, until underlying brain structure resources become so depleted that brain activity then begins to decline. Criterion B indicates that brain activity increases when a task demands more cognitive processing than an individual has available, until brain resources become depleted and again, we then see a decline in brain activity. Cabeza and Dennis (21) suggest that age affects this relationship; reaching the threshold where resources become depleted occurs earlier in older adults. The remaining two criteria describe “successful compensation”, requiring a positive correlation between hyperactivity and cognitive performance (criterion C) and a change in cognitive performance with alteration of hyperactive regions (criterion D). Criterion D suggests that if we manipulate a hyperactive region (by either disrupting it or enhancing it), we should see a coordinated decline or improvement of the associated compensatory cognitive function (21).

**Figure 1.**
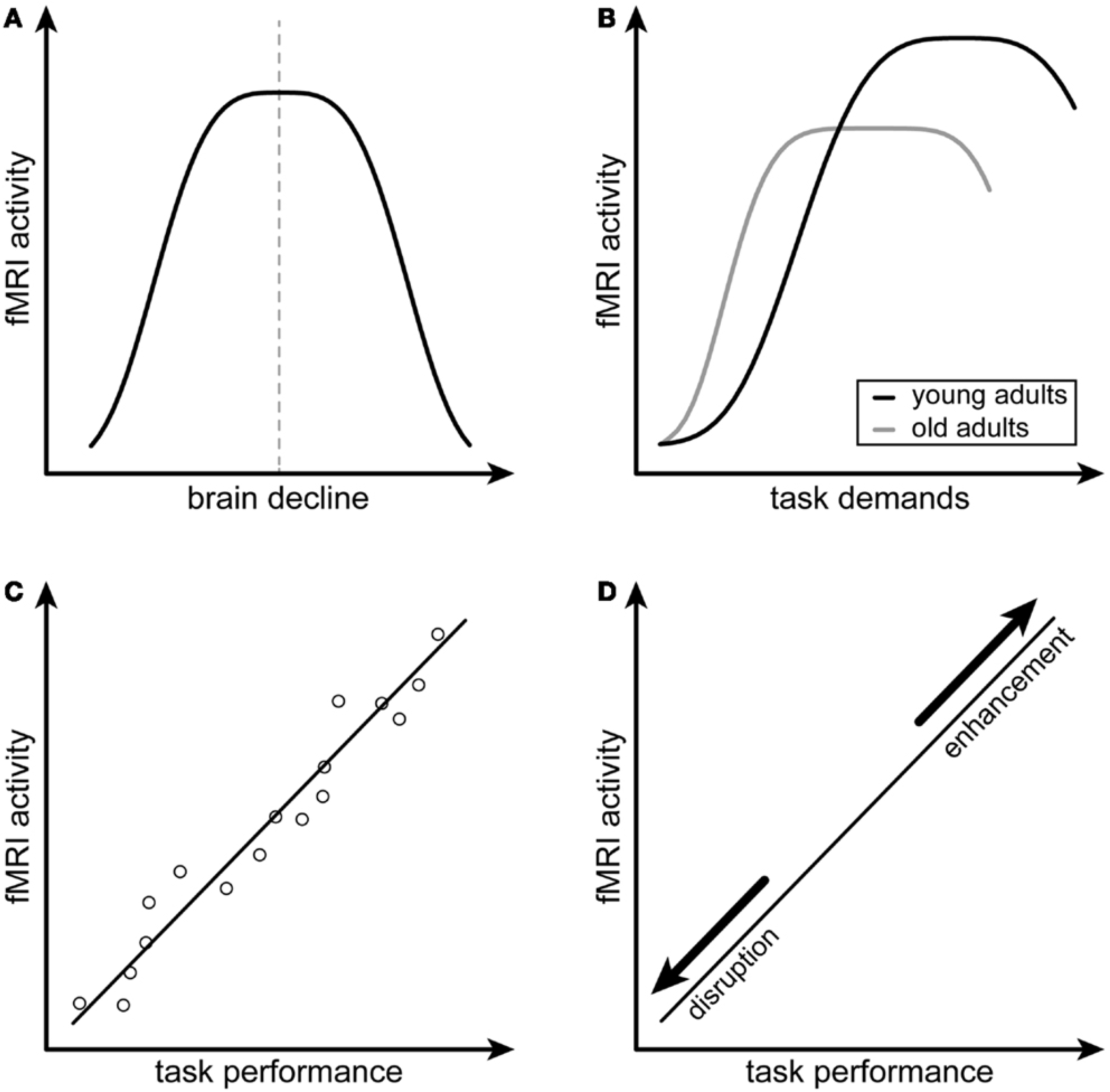
Conceptual Model for Compensatory Neural Hyperactivity by Dennis and Cabeza. (**2013**). Criterion A indicates that compensatory neural activity as measured by functional magnetic resonance imaging (fMRI), shows an inverted U-shaped relationship with brain decline. Criterion B indicates that compensatory neural activity decreases with increased task difficulty, especially in older individuals. Criterion C indicates that compensatory neural activity is positively associated with task performance. Criterion D indicates that the relationship between compensatory neural activity and task performance is disrupted or enhanced by modulating the hyperactive brain regions. Figure reprinted from Scheller, Minkova (16) under the terms of the Creative Commons Attribution License (CC BY).

Hyperactivity may explain the well-known and often controversial discrepancy between subjective and objective cognitive function in cancer survivors (13). If hyperactivity reflects neural compensation, it could mask the underlying cognitive deficit (8). However, patient awareness of the additional neural effort required to maintain performance might be reflected in low self-ratings of cognitive function compared to normal or near-normal objective cognitive performance. Therefore, determining if hyperactivity is compensatory would significantly help clarify the inconsistency between objective and subjective CRCI (22, 23) that has frequently resulted in dismissal of patient reports. Importantly, compensation-related theories suggest methods for enhancing compensation to improve cognition (15, 16). Identifying the subgroup of patients who demonstrate hyperactivity may also yield insights regarding modifiable factors that could be applied to other subgroups to help improve cognitive function.

However, compensatory hyperactivity may come at the cost of faster spread of age-related and other neuropathologies making it even more important to identify precisely which patients demonstrate this biotype. Hyperactivity may increase oxidative stress and the transfer of proteins such as tau and α-synuclein between neurons and subsequently lead to greater accumulation and aggregation (24, 25). With gliomas, hyperactivity and increased functional connectivity may increase the spread of glioma cells and impact patient survival (26). One type of neuropathology often studied in relation to CRCI is accelerated brain aging (27). Brain age is an AI-derived neuroimaging measure of brain health which when compared to chronological age yields the Brain Age Gap (BAG) (28). In our previous studies, we found that while all biotypes had higher brain age than noncancer controls, Biotype 2 (those with the best cognitive function) had lower brain age compared to the other biotypes (14, 29). However, it is unknown if neural activity and BAG are related.

To better understand hyperactivity, compensation, and brain aging, we examined neural activity in our CRCI biotypes and tested the compensatory criteria proposed by Cabeza and Dennis (21). First, we hypothesized that magnitude of neural activity differs across the CRCI biotypes and controls and is highest in Biotype 2. For compensatory criterion A, we hypothesized that hyperactivity will be related to brain decline, specifically that there would be an inverted-U relationship between gray matter volume and neural activity. For compensatory criterion B, we predicted there would be a significant negative relationship between neural activity and age, with older participants showing less compensatory hyperactivity. For compensatory criterion C, we hypothesized that neural activity would be positively correlated with cognitive performance. Given that this was a retrospective study, testing criterion D was not possible and would require a clinical trial that is beyond the scope of this study. Unrelated to compensatory criteria, we also predicted that higher neural activity would be associated with increased neuropathology (as measured by BAG).

## Methods

### Participants

We evaluated a retrospective sample (data collected between 2008-2013) of 80 breast cancer survivors and 80 noncancer, female controls. The breast cancer survivors were age 35–73 years and had completed all primary treatments (surgery, radiation, chemotherapy) excluding hormone blockade at least 6 months before study enrollment. See Table 1 for participant demographics such as age, education, time since treatment, etc. Chemotherapy regimens included doxorubicin/ cyclophosphamide (N = 3), doxorubicin/cyclophosphamide/ paclitaxel (N = 52), doxorubicin/paclitaxel (N = 1), doxorubicin/cyclophosphamide/fluorouracil (N = 1), doxorubicin/ cyclophosphamide/methotrexate (N = 5), cyclophosphamide/ paclitaxel (N = 16), and fluorouracil/epirubicin/cyclophosphamide (N = 2). Participants were free from disease and had no history of relapse or recurrence at the time of evaluation. Participants were excluded for neurologic, psychiatric, or medical conditions known to affect cognitive function. The studies involving humans were approved by Stanford University Institutional Review Board. The studies were conducted in accordance with the local legislation and institutional requirements. The participants provided their written informed consent to participate in this study.

**Table 1.**
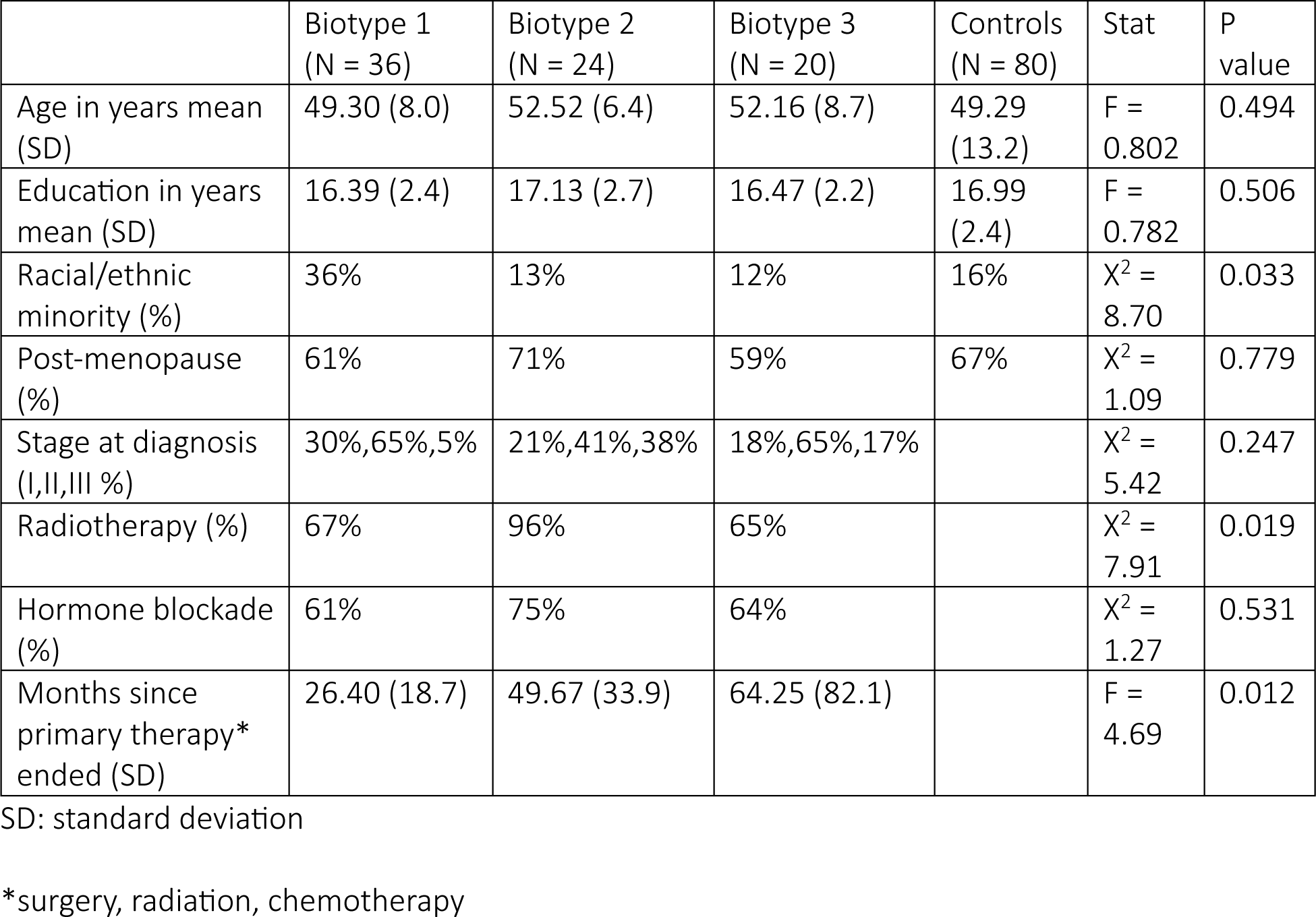
Participant Characteristics.

### Neuroimaging Data Acquisitions

Resting state fMRI data were collected using a T2* weighted gradient echo spiral pulse sequence: TR = 2000 ms, TE = 30 ms, flip angle = 80°, and 1 interleave, FOV = 22 cm, matrix = 64 × 64, in-plane resolution = 3.4375 mm2, number of volumes = 216. A high-resolution, 3D IR-prepared FSPGR anatomic MRI scan was obtained: TR=8.5, TE= minimum, flip=15 degrees, TI=400 ms, BW=+ / − 31.25 kHz, FOV=22 cm, phase FOV=0.75, slice thickness=1.5 mm, 124 slices, 256 × 256 @ 1 NEX, scan time=4:33 min. Diffusion tensor imaging data were also collected during this scan session but are not reported here. All sequences were collected using a GE Signa HDx whole body scanner (GE Medical Systems, Milwaukee, WI).

### Functional Brain Connectivity

Resting state fMRI data were preprocessed using Statistical Parametric Mapping 12 (SPM12) (30) and CONN 21a (31) implemented in Matlab v2023b (Mathworks, Inc, Natick, MA). Briefly, this involved realignment, coregistration with the segmented anatomic volume, spatial normalization, artifact detection (global signal = 3.0 standard deviations, motion = 1.0 mm, rotation = 0.05 mm), band-pass filtering (0.008–0.09 Hz), and correction of non-neuronal noise (32). Temporal correlations between all possible pairs of 268 regions (33) were computed based on the corrected fMRI signal to create a 268×268 functional connectivity matrix for each participant. Thus, the matrix describes the brain network, or connectome, comprised of nodes (regions) and edges (connections).

### Biotypes

We previously developed a machine learning algorithm for determining data-driven, latent patterns of brain abnormality (biotypes) from functional brain connectivity in this cohort. We then examined cognitive phenotypes associated with each biotype based on scores from six tests: Comprehensive Trail Making Tests 1 and 5, Delis-Kaplan Executive Function System Letter Fluency test, Immediate and Delayed Recall from the Rey Auditory Verbal Learning Test, and Global Executive Composite (GEC) of the Behavioral Rating Inventory of Executive Function Adult Version (14). Biotype 1 demonstrated impaired cognitive function on 6/6 tests, Biotype 2 had relatively preserved cognitive function with impairment on 2/6 tests, and Biotype 3 showed moderately impaired cognitive function with impairment on 4/6 tests. Impairment was defined as differing significantly from non-cancer controls (p < 0.05, corrected for multiple comparisons), although biotypes also differed significantly from each other. We then cross-validated our biotype algorithm in an independent sample (12, 14). See Table 1 for demographic and clinical details of each Biotype.

### Neural Activity

We measured neural activity from resting state fMRI using the z-transformed percent amplitude of fluctuation (zPerAF) (34). zPerAF is a measure of percent signal change and is calculated for each region as the sum of the absolute values of the standard deviation (z) normalized, mean centered signal intensities at each time point, divided by the total number of fMRI time points:

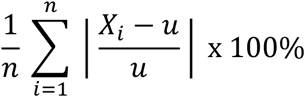

where *X_i_* = signal intensity at the *i^th^* time point, *u* = mean signal across time points, and *n* = number of time points. zPerAF, as well as mean normalized PerAF (mPerAF), have been shown to be more reliable than other metrics of resting state neural activity including ALFF and fALFF (34). We chose to utilize zPerAF given our experience that mPerAF can result in infinity values if the mean time series is zero.

### Brain Age Gap (BAG)

We estimated brain age from anatomic MRI by utilizing brainageR v2.1, a publicly available algorithm that has been shown to be one of the most reliable for predicting age from brain MRI (35). The brainageR model was trained on 3377 healthy individuals (mean age = 40.6 years, SD = 21.4, age range 18-92 years) and tested on an independent dataset of 857 healthy individuals (mean age = 40.1 years, SD = 21.8, age range 18-90 years). The model accepts raw, T1-weighted MRI scans, segments and normalizes them in SPM12 with custom templates, and utilizes the resulting gray, white, and CSF volumes in a Gaussian Processes regression to predict brain age (36, 37). Chronological age was subtracted from estimated brain age to calculate BAG, a metric of brain health wherein a positive BAG represents accelerated brain age (i.e., neuropathology), and a negative BAG represents decelerated brain age (28).

### Statistical Analysis

To test the hypothesis that magnitude of neural activity differs significantly among biotypes, we compared zPerAF between groups (biotypes and controls) using ANOVA with false discovery rate (FDR) correction for multiple comparisons. We also examined Dennis and Cabeza’s compensation criterion (Figure 1). For criterion A (inverted U-shaped relationship between fMRI activity and brain structure), we plotted zPerAF as a function of gray matter volume and fitted a polynomial regression model using these two variables. Gray matter volume was extracted from anatomic MRI using voxel-based morphometry in SPM12 (38). We then compared the polynomial model with a linear model for goodness of fit using ANOVA. We did not have fMRI task data to test compensation criterion B (compensatory hyperactivity decreases with increased task difficulty), but this criterion also indicates that older individuals show reduced compensatory hyperactivity. Therefore, we examined the Pearson correlation between zPerAF and age. To examine compensation criterion C (positive correlation between fMRI activity and task performance), we conducted Pearson correlations between zPerAF and cognitive testing scores. We did not have data to test compensation criterion D (disruption/enhancement of hyperactive brain regions alters the relationship between neural activity and task performance). To test our hypothesis that higher neural activity is associated with higher neuropathology, we conducted Pearson correlation between zPerAF and BAG.

## Results

### Neural Activity Between Groups

As shown in Figure 2a, z-normalized percent amplitude of fluctuation (zPerAF) was significantly different among biotypes and controls (p < 0.05, FDR corrected) in right temporal pole, left anterior cingulate, right inferior temporal gyrus, bilateral insular gyrus, right supramarginal gyrus, left middle frontal gyrus, left superior frontal gyrus, left inferior frontal gyrus, left medial orbital frontal gyrus, left superior medial frontal gyrus, left precentral gyrus, left superior temporal gyrus, right lingual gyrus, and right middle frontal gyrus. To reduce comparisons, we calculated the mean across these significant regions (Figure 2b) and conducted ANOVA with Tukey HSD post hoc correction (omnibus F = 13.5, p < 0.001), which indicated that Biotype 2 (n=24) showed significant hyperactivity compared to the other biotypes and controls (n=80; p < 0.004, corrected). Biotypes 1 (n=36) and 3 (n=20) showed significant hypoactivity compared to Biotype 2 and controls (p < 0.02, corrected), but were not different from each other (p = 0.931, corrected).

**Figure 2.**
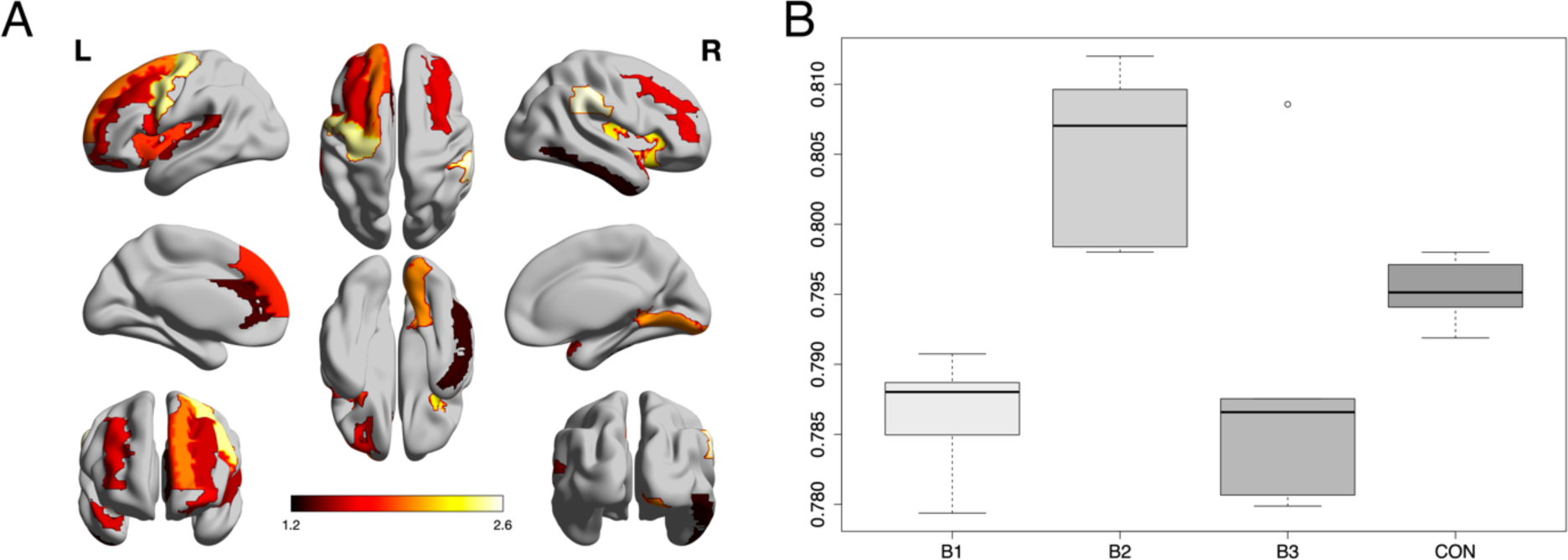
Group Differences in Neural Activity. A: Z-normalized percent amplitude of fluctuation (zPerAF) was significantly different among biotypes and controls (p < 0.05, FDR corrected) in right temporal pole, left anterior cingulate, right inferior temporal gyrus, bilateral insular gyrus, right supramarginal gyrus, left middle frontal gyrus, left superior frontal gyrus, left inferior frontal gyrus, left medial orbital frontal gyrus, left superior medial frontal gyrus, left precentral gyrus, left superior temporal gyrus, right lingual gyrus, and right middle frontal gyrus. Color bar indicates the log of the p value. B: The mean zPerAF across significant regions for each group is displayed as a boxplot. Biotype 2 (B2) showed significant (p < 0.004, corrected) hyperactivity compared to the other biotypes and controls. Biotypes 1 and 3 showed significant (p < 0.02, corrected) hypoactivity compared to Biotype 2 and controls.

### Compensation Criterion A

Given that only Biotype 2 showed hyperactivity, compensation criterion analyses were performed only in this group. The scatterplot of zPerAF as a function of gray matter volume indicated the expected inverted U-shaped relationship (Figure 3). We used the mean zPerAF across significant regions to reduce comparisons in this small sample. The polynomial fit was significant (R^2^ = 0.42, p = 0.029) including the polynomial term (p < 0.010). The linear fit was not significant (R^2^ = 0.01, p = 0.776) and was a significantly poorer fit of the data compared to the polynomial model (F = 9.2, p = 0.010).

**Figure 3.**
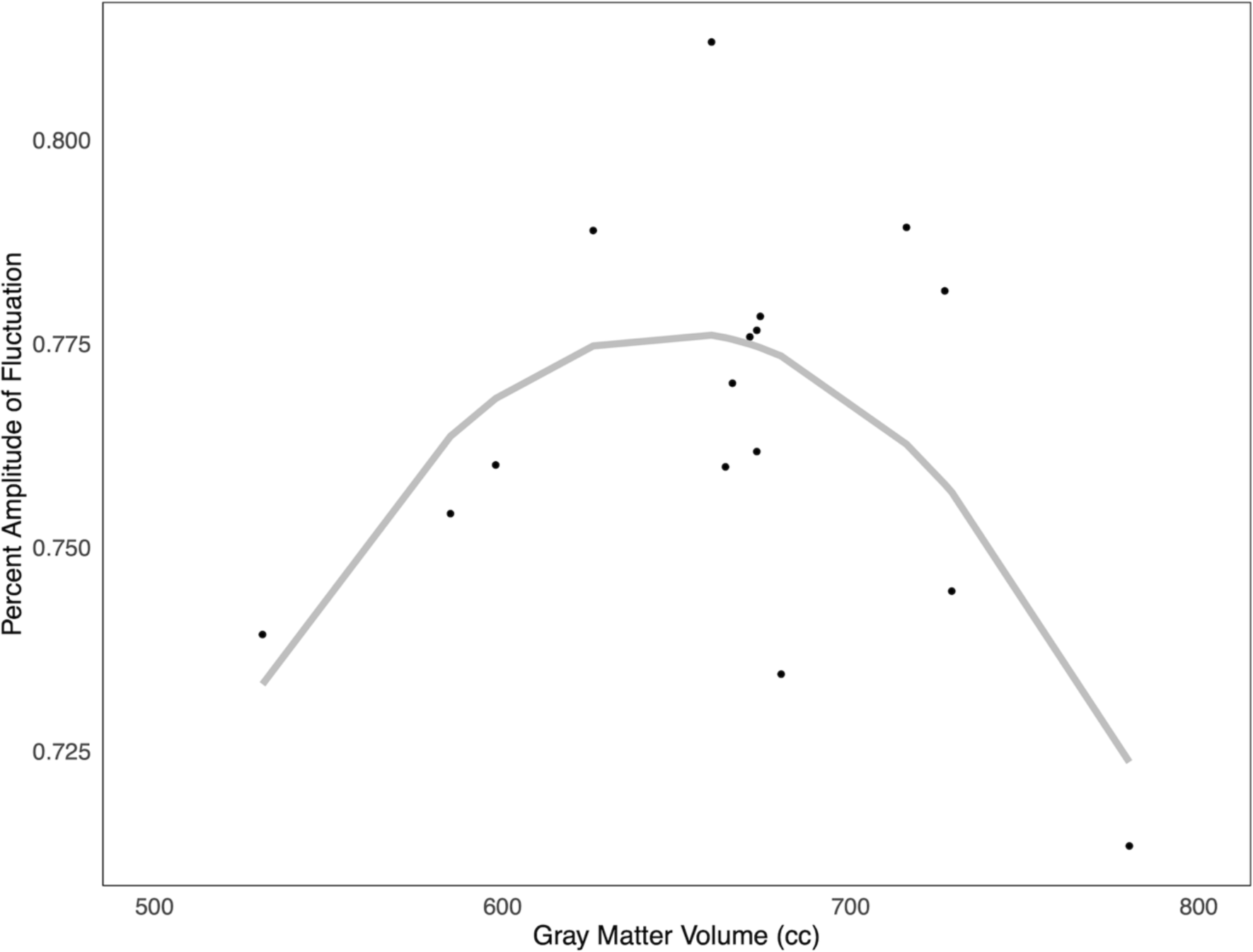
Compensation Criterion A. In Biotype 2, there was an inverted U-shaped relationship between z-normalized percent amplitude of fluctuation (zPerAF) and gray matter volumes consistent with Dennis and Cabeza’s criterion A for compensatory neural hyperactivity. These data showed goodness of fit with a polynomial (R^2^ = 0.42, p = 0.029) but not a linear model (R^2^ = 0.01, p = 0.776).

### Compensation Criterion B (Partial)

Again, without task fMRI data, we were only able to examine the part of this criterion related to age. We observed the expected negative relationship between age and mean zPerAF across significant regions in Biotype 2 (r =-0.53, p = 0.018).

### Compensation Criterion C

To reduce comparisons, we used the mean zPerAF across significant regions and evaluated only the cognitive tests that we previously showed to be significantly different in Biotype 2 compared to the other biotypes or controls (14). Consistent with criterion C, members of Biotype 2 showed significant positive correlations between Letter Fluency test score and zPerAF (r = 0.68, p = 0.004) as well as GEC and zPerAF (r = 0.566, p = 0.011). Both correlations indicated better cognitive function with higher neural activity.

### Neural Activity and Brain Age Gap

Although we previously found that Biotype 2 had the lowest brain age of the biotypes (12), correlation results between mean zPerAF and BAG indicate that members of Biotype 2 with the highest neural activity have accelerated brain aging (r = 0.45, p = 0.042).

## Discussion

Here we found that neural activity differed across our three CRCI Biotypes and healthy controls, primarily in prefrontal cortex. The regions included the right temporal pole, left anterior cingulate, right inferior temporal gyrus, bilateral insular gyrus, right supramarginal gyrus, left middle frontal gyrus, left superior frontal gyrus, left inferior frontal gyrus, left medial orbital frontal gyrus, left superior medial frontal gyrus, left precentral gyrus, left superior temporal gyrus, right lingual gyrus, and right middle frontal gyrus. As predicted, Biotype 2 demonstrated significant hyperactivity in these regions compared to the other biotypes and controls.

Further examination of Biotype 2 showed that this hyperactivity met several of the criteria to be considered compensatory (21). Regarding criterion A, there was an inverted U-shaped relationship between brain activity and gray matter volume. Although we did not have a measure of task demand to fully test criterion B, we found the expected negative relationship between age and neural activity, with activity decreasing with older age. Next, for criterion C, we found a positive relationship between neural activity and cognitive performance, with hyperactivity being associated with better performance on an objective cognitive test as well as with higher self-ratings of cognitive function. This provides evidence of successful compensatory brain activity, but also suggests that patients may not be aware of the additional neural effort required to preserve their cognitive function as we expected. Self-assessment of cognitive function may need to occur closer to objective cognitive loading tasks to evaluate this relationship more precisely. As noted above, we were unable to test criterion D. This would require a behavioral or pharmacologic trial to examine the mediating effect of disruption/enhancement of hyperactive brain regions on the relationship between neural activity and task performance.

This was the first study of CRCI to identify which patients show neural hyperactivity. Identifying which subgroup of patients demonstrates a specific disease biomarker is essential for precision medicine given that different subgroups will likely have different responses to various interventions. Our results reveal a specific mechanism (prefrontal activity) that may result in CRCI in different groups of patients, which could help determine which treatments and prevention strategies will be most effective for each patient. As we reported previously, there were no distinguishing demographic or clinical characteristics of Biotype 2 expression that could explain their relatively preserved cognitive function (12, 14). Our present results suggest that prefrontal hyperactivity may be responsible for this difference in outcome compared to other patients.

Accordingly, our study was also the first to explicitly test that neural hyperactivity meets the criteria to be considered compensatory (39–41). Our findings are in line with what has been reported as compensatory activity during aging (21). This is relevant because many studies show that CRCI may reflect age acceleration (42–47). Cabeza and Dennis (21) showed that age-related increases in functional connectivity met three of the four compensatory criteria; (A) increased functional activity in the frontal cortex during healthy aging and mild cognitive impairment but decreased functional activity during more severe impairments; (B) examinations of memory load showed that frontal cortex connectivity has an inverted-U relationship with task demand; (C) age-related increases in frontal cortex functional connectivity was related to successful cognitive performance. Other studies show similar relationships between age and brain activity as well as brain activity and cognitive performance and task demand (48–53). High performing older adults (demonstrating preserved cognitive function, similar to CRCI Biotype 2) show increased frontal cortex activity, compared to low performing older adults (demonstrating impaired cognitive function, similar to CRCI Biotype 1) (48–51).

It remains unclear why or how Biotype 2 patients demonstrate compensatory neural hyperactivity. Given the retrospective nature of our studies, we likely lack the data necessary to determine what sets them apart from other patients. It will be essential to conduct prospective biotyping studies to determine if there are modifiable factors contributing to the cognitive phenotype of Biotype 2. However, while compensatory hyperactivity in Biotype 2 may help explain their increased cognitive resilience, they could also be at risk for accelerated brain aging. Our results showed a positive relationship between neural activity and brain age gap (BAG, a proxy of neuropathology). In a study of healthy adults, Scheller et al. (53) found that together APOE variant and brain age moderated the relationship between neural hyperactivity and cognitive performance. Specifically, APOEe4 carriers with higher brain ages had increased frontal cortex activity which correlated with preserved cognitive function (53). Unfortunately, we cannot determine the directional nature of this relationship in either study, as data was collected at a single time point. Neurodegeneration may result in hyperactivity, or this relationship could be bidirectional. In the current study, given that 1) hyperactivity was observed only in Biotype 2 and met several criteria for being compensatory and, 2) these patients demonstrated a unique relationship between BAG and hyperactivity while simultaneously having the lowest BAG, it is more likely that hyperactivity in this subgroup results in neurodegeneration rather than the reverse. However, further studies are required to better evaluate these relationships.

Previous studies of cognitive impairment in aging adults found a relationship between accelerated brain aging, worsening cognitive function, and clinical disease severity (54–56). Brain age at baseline predicted a future advancement from mild cognitive impairment to Alzheimer’s disease three years later, and this data was used to create hazard ratios for the development of Alzheimer’s based on brain age (54–56). Future studies should include repeated brain imaging and cognitive testing for cancer survivors to determine if compensatory activity precedes increases in brain age, or if brain age can predict further future cognitive declines in those with CRCI.

Our results provide novel insights regarding potential interventions for CRCI by identifying *who* has hyperactivity and *where* hyperactivity occurs. Methods for enhancing compensation to improve cognition include neuromodulation (15, 16). Neuromodulation is a strong candidate for addressing abnormal neural activity as it is already FDA approved for use in other neuropsychiatric conditions (57). Future prospective studies of neural hyperactivity could determine which patients might benefit most from such strategies, including some of the potential risks (brain aging) and benefits (compensatory cognition) of up versus downregulation, respectively.

This study is not without limitations. As mentioned previously, as this was a retrospective study, we did not have a measure of task demand to be able to fully test Criterion B. Future prospective studies should include measures of task demand when studying CRCI; for example, dual- or concurrent-tasks, tasks that vary demand, linguistic analyses, physiological measures, or self-report measures such as the NASA Task Load Index (58–65). Including self-report measures of cognitive load or demand after each objective cognitive test could also assess whether patients are aware of any increased neural effort associated with their performance. In addition, interventions which target the hyperactive brain regions could be examined to directly test Criterion D. For example, researchers could utilize methods of brain stimulation (e.g., transcranial magnetic stimulation, transcranial alternating current stimulation, transcranial direct current stimulation) or neurofeedback with CRCI patients to examine the effect of these noninvasive brain manipulations on cognition (e.g., 66, 67-72). In addition, animal models could be used to examine the effects of direct electrical stimulation of brain regions linked to CRCI. Both BAG and zPerAF are measured from neuroimaging and although they are derived from different imaging modalities, there is inherent neurobiological overlap. Therefore, future studies should examine the effect of neural activity on non-imaging biomarkers of neurodegeneration such as peripheral tau and amyloid-beta (73), for example. Finally, this study only includes breast cancer survivors, and those who have undergone treatment for other cancer types with other treatment regimens may differ in brain activity post-treatment and may not display the same compensatory mechanisms.

Overall, the current study demonstrates that the neural hyperactivity observed in CRCI Biotype 2 potentially meets most of the compensatory criteria. This neural compensation may explain the preserved cognitive function observed in Biotype 2 compared to the other CRCI Biotypes. Further, neural hyperactivity may be related to accelerated brain aging. Future studies should include measures of cognitive decline and manipulation of frontal cortex activity to further test the compensatory criteria, as well as collection of longitudinal data to further elucidate the relationship between hyperactivity and brain aging.

## Acknowledgements

The authors would like the thank the faculty and staff of the Richard M. Lucas Center at Stanford University. This research was supported by the National Institutes of Health (AG078411 to MMM and R01CA172145, R01CA226080, and DP2OD004445 to SRK). The sponsor was not involved in any aspects of this study.

## Author contributions

Conceptualization: MMM, SRK. Data collection: SRK, DB. Formal analysis: ADS, SRK. Methodology: All authors. Writing-review & editing: All authors.

## Data Availability

All data relevant to the study are included in the article. The original MRI data underlying this article cannot be shared publicly due to data protection regulation.

## Additional Information

The authors declare no competing interests related to this manuscript.

